# Preliminary Findings from an Augmented Reality (AR) App Delivering Recovery-Oriented Cognitive Therapy for Negative Symptoms in Schizophrenia

**DOI:** 10.1101/2024.10.28.24315453

**Authors:** Sunny X. Tang, Moein Foroughi, Aaron P. Brinen, Michael L. Birnbaum, Sarah Berretta, Leily M. Behbehani, John M. Kane, Edward Yoon, William F. Cronin

**Affiliations:** North Shore Therapeutics, Inc., Rochester, MN.; Feinstein Institutes for Medical Research, Northwell Health, Manhasset, NY; Zucker Hillside Hospital, Northwell Health, Glen Oaks, NY; Donald and Barbara Zucker School of Medicine, Hempstead, NY; Vanderbilt University Medical Center, Nashville, TN; Columbia University Vagelos College of Physicians and Surgeons, New York, NY; New York State Psychiatric Institute, New York, NY; School of Behavioral and Brain Sciences, University of Texas at Dallas, Richardson, TX; Department of Psychology, Yale University, New Haven, CT

**Author notes:** **Corresponding Author:** Sunny X. Tang, M.D., North Shore Therapeutics, Inc., 2423 Virginia Ln. SW, Rochester, MN 55902.

**Keywords:** Schizophrenia, negative symptoms, augmented reality, CBT, digital health, app, CT-R

## Abstract

**Background:** Negative symptoms are a primary driver of poor outcomes in schizophrenia spectrum disorders (SSD), but there are no FDA-approved medications or FDA-cleared therapeutics targeting negative symptoms in schizophrenia. NST-SPARK is a novel digital therapeutic targeting negative symptoms in SSD. It is a smartphone application delivering recovery-oriented cognitive therapy (CT-R), via gamified augmented reality (AR) experiences, to provide experiential learning aimed at dismantling maladaptive beliefs. In this study, we assessed a prototype (NST-SPARK v.1.5) in 20 participants with SSD and clinically significant negative symptoms. NST-SPARK v.1.5 delivers a single therapeutic module over a 1-week period. The primary objective was to determine the acceptability and feasibility of this approach. Secondary objectives were to generate descriptive findings for changes in defeatist beliefs, self-esteem, and attitudes toward goal-oriented activities.

**Methods:** Recruitment and all study procedures were completed online. Twenty participants with schizophrenia or schizoaffective disorder were enrolled, with a range of demographic and socioeconomic status and treatment settings. Participants completed self-reports on the acceptability and feasibility of NST-SPARK v.1.5 and provided open-ended feedback through a semi-structured interview. Self-report scales on defeatist beliefs, self-esteem, and attitudes toward goal-oriented activities were completed before and after participants were introduced to NST-SPARK, and then again at a 1-week follow-up.

**Results:** In general, participants found NST-SPARK v.1.5 to be feasible and acceptable, responding with an average response of “Agree”, indicating that the intervention was found to meet with the participants’ approval and seemed implementable. Almost all participants (19 of 20) used the app on their own prior to the 1-week follow up despite not being incentivized to do so. In addition, participants responded to open-ended feedback questions in a generally positive way. We also observed shifts in defeatist beliefs (Cohen’s d = 0.12), self-esteem (Cohen’s d = -0.21), and attitudes toward goal attainment consistent (intention: Cohen’s d = 0.13; confidence: Cohen’s d = 0.35) with intended improvements in these targeted areas. Participants were able to make substantive progress toward identified goals in 90% of cases.

**Conclusions:** This preliminary, single-arm, unblinded study of a single-module prototype for NST-SPARK found that the approach is generally acceptable and feasible for people with SSD and negative symptoms. Engagement of the intended target of defeatist beliefs was supported by our findings but require confirmation in future randomized controlled trials. Overall, NST-SPARK is based on a promising approach and further development is warranted.

## Introduction

Effective treatment for negative symptoms in schizophrenia spectrum disorders (SSD) is a major unmet need. SSD are among the most devastating psychiatric disorders. Onset typically occurs in late adolescence or early adulthood and can severely disrupt completion of educational goals [1], obtaining and sustaining employment, and developing healthy relationships [2,3]. While there are effective treatment options for positive psychotic symptoms, negative symptoms such as avolition, anhedonia, asociality, and emotional flattening typically persist despite evidence-based care [4,5]. In a meta-analysis including over 12,000 individuals and 168 randomized-controlled trials of interventions for negative symptoms in schizophrenia, few statistically significant effects on negative symptoms were evident, and none reached the threshold for clinically meaningful improvement [6]. Prominent clinically significant negative symptoms are experienced by up to 60% of individuals with SSD and are often considered to be the greatest contributors to functional disability [7]. Among the negative symptoms, avolition appears to be at the core, driving the greatest share of functional impairment and perhaps driving other negative symptoms [8,9]. Currently, there are no FDA-approved medications or FDA-cleared therapeutics specifically targeting avolition or any of the negative symptoms of schizophrenia.

NST-SPARK is a novel digital therapeutic targeting negative symptoms. It is designed as a smartphone application that delivers recovery-oriented cognitive therapy (CT-R), via gamified augmented reality (AR) experiences, to provide experiential learning aimed at dismantling maladaptive beliefs.

CT-R is an evidence-based psychotherapy with efficacy in reducing avolition specifically, negative symptoms in general, and in improving functioning among individuals with SSD [10,11]. In contrast to CBT for psychosis, CT-R prioritizes negative symptoms by focusing on patient engagement, correcting defeatist beliefs, and reinforcing positive, recovery-oriented beliefs and actions [12,13]. It places greater emphasis on empowerment and engagement, and specifically targets avolition with high fidelity. Defeatist performance beliefs are overgeneralized negative thoughts about one’s ability to accomplish tasks; they have been related to in-the-moment negative symptoms assessed via ecological momentary assessment [14] and contribute to the development and maintenance of negative symptoms. CT-R attempts to correct defeatist beliefs by engaging individuals in rewarding activities while activating an adaptive mode of functioning. A randomized controlled trial comparing CT-R with standard treatment for 18 months in patients with SSD found significant reductions in avolition-apathy negative symptoms (Cohen’s d = -0.66, p = 0.01) and improvement in global functioning (Cohen’s d = 0.56, p = 0.03) compared to treatment as usual [11]. In a follow up randomized controlled trial, between-group differences for negative symptoms were evident after 6 months of treatment, and benefits persisted 6 months after treatment was concluded [10]. However, while promising, CT-R is limited by its reliance on highly trained clinicians. To maximize quality and access to care, clinicians need effective and customizable resources that can be used to elicit, identify, and treat dysfunctional thought patterns and behaviors contributing to negative symptoms in SSD.

Immersive technologies have been deemed promising, safe, and feasible for patients with SSD [15,16], with no significant adverse effects demonstrated in a recent meta-analysis on AR and VR interventions. AR was chosen for implementation of CT-R because it allows for delivery of an engaging experience while remaining anchored in the “real world.” This unique characteristic of AR was intended to encourage patients to interact with their surroundings instead of retreating to a simulated space, thereby potentially increasing the translation of lessons learned to the “real world” where they are needed [17]. Unlike virtual reality (VR), which requires specialized hardware, AR can be implemented on smartphones, which increases its real-world usability and scalability, and decreases implementation costs, an established barrier to commercialization of several prior products. Meanwhile, it delivers a stimulating immersive experience that can interrupt negative feedback loops that perpetuate negative symptoms by transporting the patient to a novel environment. The simulated experiences in AR can be used to engage reward circuitry and motivate users to change behavior by incorporating elements of gaming [18]. Gamification involves using game-based elements and game thinking to motivate action and promote learning and engagement. The global phenomenon Pokémon Go, for example, demonstrated an increase in physical activity and a decrease in psychological distress in non-psychiatric populations [19,20]. The popular language learning app, Duolingo, uses gamification elements in language learning and has shown to positively affect learners’ behavior, commitment, and motivation [21]. Game-like digital environments hold promise to improve negative symptoms through activating reward circuitry and eliciting positive experiences and emotions in people with SSD by leveraging proven design elements like points, rewards and progress tracking [22,23], thereby creating a neuropsychological state that may be conducive to treatment.

In this study, we assessed a prototype of NST-SPARK (NST-SPARK v.1.5) in 20 volunteers with SSD and clinically significant negative symptoms. NST-SPARK v.1.5 delivers a single session of CT-R, along with two AR experiences. Participants used the app over a 1-week period. The primary objective was to determine the acceptability and feasibility of this approach. Secondary objectives were to generate descriptive findings for changes in defeatist beliefs, self-esteem, and attitudes toward goal-oriented activities.

## Methods

### Participants

Recruitment was completed from volunteers from prior market research studies as well as online through referrals and social media advertisements (Reddit, Twitter, and Facebook). All research procedures were done remotely, through video conferencing, by two centralized trained assessors. Interested potential participants completed an initial screening visit to determine eligibility. Diagnoses were confirmed by the Structured Clinical Interview for DSM-5 (SCID-5) conducted at the screening visit [24]. Additional inclusion criteria included age 18-65 years, all genders, races, and ethnicities, dependable access to stable Wi-Fi and a device with videoconferencing ability, able to provide informed consent, proficient in English, and clinically significant negative symptoms (rating of 2 or higher on the Avolition/Apathy or Anhedonia/Asociality subscale of the Scale for the Assessment of Negative Symptoms (SANS) [25]. Exclusion criteria included acute safety concerns, recent change in level of care (medication changes within 4 weeks or acute psychiatric care within 12 weeks), substance-induced psychotic disorder, or significant cognitive or physical limitations that prevented the participant from being able to operate NST-SPARK. Eligible participants provided informed consent via a secure digital platform, Zoho. All study procedures were approved by the BRANY IRB #23-02-339-1483. This study is registered on ClinicalTrials.gov #NCT06653829.

Twenty participants with schizophrenia or schizoaffective disorder were enrolled, with a range of demographic and socioeconomic status and treatment settings (Table 1). In total, participants completed a screening visit, followed by a Visit 1 where they were introduced to the app, and then a 1-week follow-up at Visit 2. Compensation was provided at study conclusion; participants had the choice between keeping the study iPhone (a second-generation iPhone SE) or sending it back for a $100 gift card of their choice.

**Table 1:**
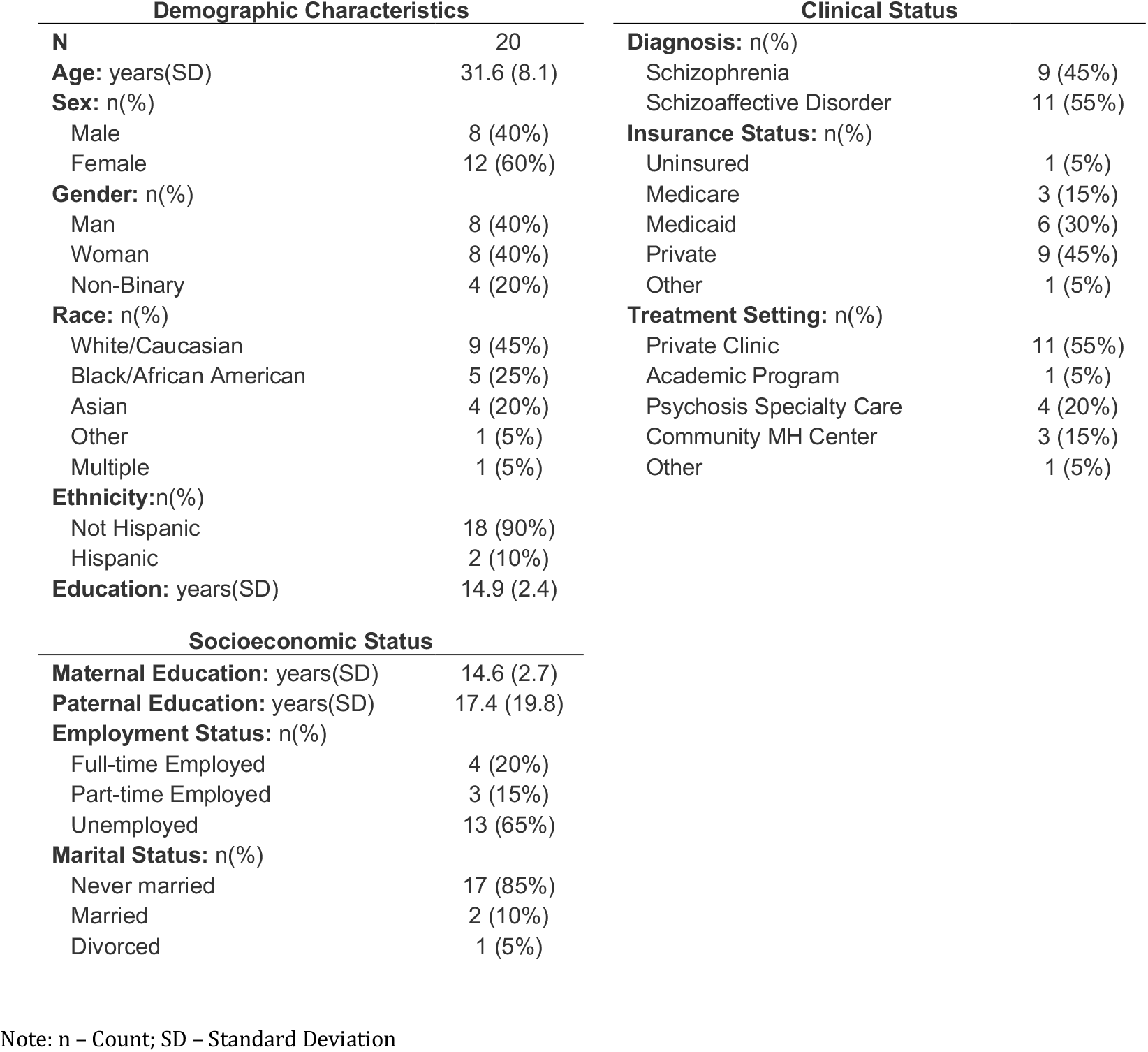
Participant Characteristics.

### NST-SPARK v.1.5

Participants received an unlocked second-generation iPhone SE with the NST-SPARK v.1.5 application. Upon receiving the iPhone, participants completed Visit 1 on Zoom with a research team member, where they were introduced to the application. NST-SPARK v1.5 delivers a single therapeutic session based on CT-R (Fig 1). The app leads participants to (A/B) elicit targeted defeatist beliefs, (C) engage in a brief (1-2min) gamified AR experience where participants received encouraging prompts as they attempted to sort flying objects based on their color, (D) draw conclusions based on the experience, and (E) generalize the lesson to positive real-life pursuits. A second AR experience was also included, interspersed with CT-R based prompts. In the second AR experience, cartoon visual stimuli (bottles) were placed around the AR space. Participants would move around their physical space to collect the bottles. In total, NST-SPARK v.1.5 took 10-15 min to complete.

**Figure 1.**
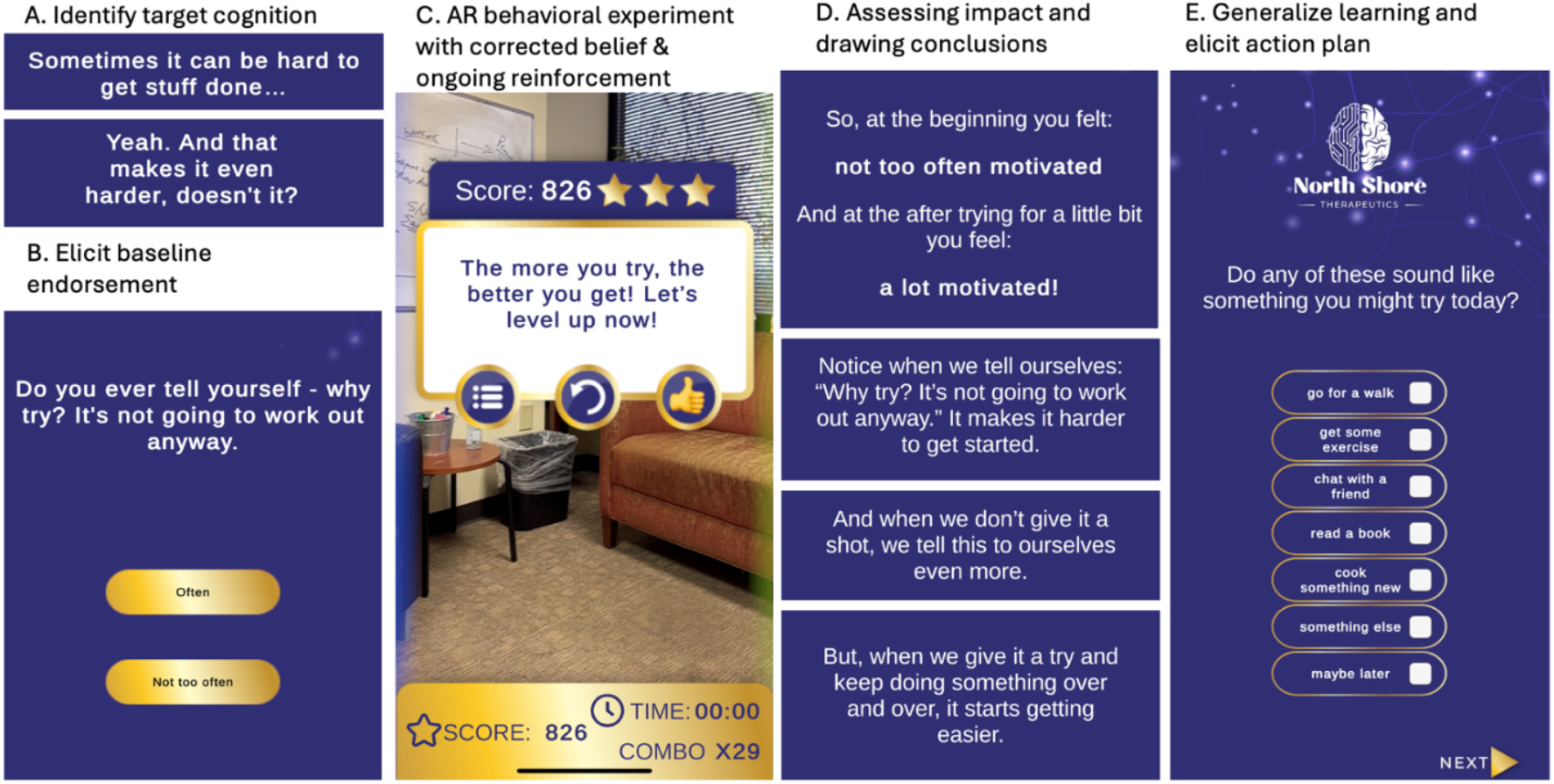
NST-SPARK v.1.5 User Experience.

### Assessments

At Visit 1, participants completed self-report questionnaires both before and after completing NST-SPARK. Questionnaires included the Defeatist Belief Scale (DBS) [26], Beck Self-Esteem Scale (BSES) [27], followed by a custom scale for Attitudes Toward Behavioral Change (NST-ATT; Appendix A). Briefly, NST-ATT asked participants to identify two tasks or goals they wanted to accomplish, one that’s relatively easy and achievable (Task A) and one that is very important to them (Task B). Participants then rated their level of intention to accomplish the task, and their confidence in being able to do so. Participants were then prompted to launch NST-SPARK v.1.5 on their study iPhone and to complete the two AR activities while the research assistant was available on Zoom to provide technical guidance. Upon completion of the NST-SPARK session, participants repeated the DBS, BSES, and NST-ATT. In addition, participants responded to the Acceptability of Intervention Measure (AIM) and Feasibility of Intervention Measure (FIM) self-report scales [28], as well as a semi-structured interview eliciting feedback on their experience with NST-SPARK v.1.5 (Appendix B).

Participants were given the recommendation of repeating NST-SPARK in the intervening week before Visit 2, but there was no incentive given for doing so. Research assistants also checked in with participants to help navigate technical issues during the intervening period. Visit 2 occurred 1 week after participants were introduced to NST-SPARK. Visit 2 began with a semi-structured interview regarding their experiences with the app over the last week (Appendix B). Participants were then asked to repeat the DBS and the BSES, as well as report on any progress made toward the tasks identified during Visit 1 on the NST-ATT.

### Statistical Analysis

As this is a preliminary study with the primary objective of determining the acceptability and feasibility of NST-SPARK v.1.5. The results are primarily represented with straightforward descriptive statistics (mean, standard deviation [SD], percentages), as well as quotes and qualitative descriptions from open-ended participant feedback. We also report Cohen’s *d* effect sizes for changes in the exploratory outcomes on defeatist beliefs, self-esteem, and attitudes toward goal-oriented activities. All analyses were done in R v.4.2.0 [29].

## Results

### Acceptability and Feasibility of NST-SPARK v.1.5

In general, participants found NST-SPARK v.1.5 to be feasible and acceptable, responding with an average response of “Agree” to the AIM and FIM questionnaires, indicating that the intervention was found to meet with the participants’ approval and seemed implementable. Table 2 summarizes the participants’ responses to the AIM and FIM questionnaires. Almost all participants (19 of 20) used the app on their own prior to the 1-week follow up.

**Table 2:** Self-Report Responses for Acceptability and Feasibility of NST-SPARK.

|  | **Visit 1 - Timepoint 2**<br>**After NST-SPARK** | **Effect Size**<br>**T2 - T3** | **Visit 2 - Timepoint 3**<br>**1-week Follow Up** |
| --- | --- | --- | --- |
| NST-SPARK meets my approval. | 3.9 (0.6) | 0.07 | 3.9 (0.9) |
| NST-SPARK is appealing to me. | 3.6 (1.0) | 0.27 | 3.9 (0.8) |
| I like NST-SPARK. | 3.8 (0.7) | 0.06 | 3.9 (0.9) |
| I welcome NST-SPARK. | 3.9 (0.7) | 0.51 | 4.2 (0.7) |
| Mean AIM Score | 3.8 (0.6) | 0.26 | 4.0 (0.7) |
| NST-SPARK seems implementable. | 3.8 (0.9) | 0.18 | 3.9 (0.8) |
| NST-SPARK seems possible. | 4.1 (0.6) | 0.07 | 4.2 (0.9) |
| NST-SPARK seems doable. | 3.8 (0.9) | 0.28 | 4.0 (0.9) |
| NST-SPARK seems easy to use. | 3.5 (1.0) | 0.37 | 3.9 (0.9) |
| Mean FIM Score | 3.8 (0.7) | 0.30 | 4.0 (0.6) |

*Note: AIM – Acceptability of Intervention Measure; FIM – Feasibility of Intervention Measure*

In addition, the participants responded to open-ended feedback questions in a generally positive way. Overall, the approach was found to be acceptable, with the main limitations identified in the mechanisms of the NST-SPARK v.1.5 prototype. Their first reactions to the app ranged from largely positive related to the gamification (“sparkly theme” “It’s pretty cool, I like that it was interactive.” “I really liked actual activity”) and technology (“I liked the AR”) to some negative reactions related to program bugs (“the tapping didn’t work”) and not knowing the purpose of the app. When asked what they liked most, many responded to the interactive nature (“It says things to validate what you are thinking”), the therapeutic message (“I liked the idea of creating energy through action. The basic idea of it was fun.”), and the sense it helped (“it helped me get out of bed which was so nice”). Most responses to what was liked least revolved around user interfaces (“the second level was too long”) and one participant brought up concerns related to accessibility for people with mobility issues. When asked if they would use it if prescribed by a professional, largely participants indicated they would use it (18 of 20): some with enthusiasm (“Yes. I think it could reach a point where you’d want to play it three or more times a week”), some due to power of the prescriber (“Yes, because it is not that challenging. I trust my doctor.”), and a small proportion of participants indicated they would not be willing to use the app because they did not find it useful (2 of 20 participants).

### NST-SPARK Effects on Defeatist Beliefs, Self-Esteem, and Goal Attainment

We observed shifts in attitudes toward goal attainment, defeatist beliefs and self-esteem in the intended directions. Overall change in defeatist beliefs was small (Table 3), with total score on the DBS remaining stable before and after using NST-SPARK during Visit 1, and then improving slightly at the 1-week follow-up (Cohen’s d = 0.12 relative to initial assessment). The largest shifts occurred in “If I do not do as well all the time, people will not respect me” (Cohen’s d = 0.23) and “If I fail partly, it is as bad as being a complete failure” (Cohen’s d = 0.21).

**Table 3:** Changes in Defeatist Beliefs.

|  | Visit 1 - Timepoint 1<br>Before NST-SPARK | Effect Size<br>T1 - T2 | Visit 1 - Timepoint 2<br>After NST-SPARK | Effect Size<br>T2 - T3 | Visit 2 - Timepoint 3<br>1-week Follow Up | Effect Size<br>T1 - T3 |
| --- | --- | --- | --- | --- | --- | --- |
| If I do not do as well all the time, people will not respect me. | 3.4 (1.8) | 0.08 | 3.5 (1.7) | 0.15 | 3.8 (1.6) | 0.23 |
| I cannot be happy unless most people I know admire me. | 4.6 (1.6) | -0.17 | 4.3 (1.9) | 0.06 | 4.4 (1.5) | -0.13 |
| If I do not do as well as other people, it means I am an inferior human being. | 4.5 (1.8) | 0.14 | 4.7 (1.8) | -0.17 | 4.4 (1.7) | -0.03 |
| If you cannot do something well, there is little point in doing it at all. | 4.8 (1.9) | 0.03 | 4.8 (1.9) | 0.15 | 5.1 (1.4) | 0.18 |
| If I fail partly, it is as bad as being a complete failure | 4.7 (1.8) | -0.09 | 4.5 (1.6) | 0.31 | 5.0 (1.6) | 0.21 |
| DBS Total Score | 21.9 (6.6) | 0.00 | 21.9 (8.2) | 0.11 | 22.7 (6.4) | 0.12 |

Note: Participants indicated their agreement or disagreement with the statements shown. Higher scores represent *disagreement* with the statements, i.e., *lower* defeatist beliefs. For each timepoint, the mean score with deviation is shown. Effect sizes are calculated with the formula for Cohen's d. *DBS – Defeatist Beliefs Scale*.

There were larger effect sizes for self-esteem (Table 4), with improvements in the total score for BSES before and after using NST-SPARK (Cohen’s d = -0.09) and then again at the 1-week follow-up (Cohen’s d = -0.12), for an overall shift of effect size -0.21. The largest overall shifts occurred in the endorsement of being “Desirable / Undesirable” (Cohen’s d = -0.55) and being “Successful / Unsuccessful” (Cohen’s d = -0.37).

**Table 4:** Changes in Self-Esteem.

|  | Visit 1 - Timepoint 1 | Effect Size | Visit 1 - Timepoint 2 | Effect Size | Visit 2 - Timepoint 3 | Effect Size |
| --- | --- | --- | --- | --- | --- | --- |
|  | Before NST-SPARK | T1 - T2 | After NST-SPARK | T2 - T3 | 1-week Follow Up | T1 - T3 |
| Successful / Unsuccessful | 5.9 (2.2) | -0.41 | 5.0 (2.2) | 0.00 | 5.0 (2.6) | -0.37 |
| Attractive / Unattractive | 5.8 (2.4) | 0.10 | 6.1 (2.5) | -0.24 | 5.5 (2.6) | -0.14 |
| Popular / Unpopular | 7.1 (2.6) | -0.13 | 6.8 (2.6) | 0.02 | 6.8 (3.1) | -0.11 |
| Independent / Dependent | 5.1 (2.7) | -0.08 | 4.9 (2.3) | -0.06 | 4.8 (2.4) | -0.14 |
| Honest / Dishonest | 3.8 (2.8) | -0.09 | 3.5 (2.5) | -0.24 | 3.0 (2.1) | -0.32 |
| Desirable / Undesirable | 6.5 (2.2) | -0.37 | 5.7 (2.1) | -0.19 | 5.3 (2.1) | -0.55 |
| Strong / Weak | 5.0 (2.4) | 0.06 | 5.2 (2.4) | -0.28 | 4.5 (2.2) | -0.22 |
| Smart / Dumb | 4.1 (2.0) | 0.00 | 4.1 (2.6) | 0.00 | 4.1 (2.4) | 0.00 |
| Powerful / Powerless | 5.4 (2.1) | 0.00 | 5.4 (2.2) | 0.04 | 5.5 (2.5) | 0.04 |
| Lovable / Unlovable | 5.0 (2.4) | 0.06 | 5.1 (2.3) | -0.18 | 4.7 (2.1) | -0.11 |
| Pleasant / Unpleasant | 4.2 (2.4) | -0.08 | 4.0 (2.4) | -0.17 | 3.6 (2.3) | -0.26 |
| Efficient / Inefficient | 4.7 (2.3) | 0.14 | 5.0 (2.8) | -0.05 | 4.9 (2.8) | 0.08 |
| Responsible / Irresponsible | 4.5 (2.4) | -0.17 | 4.2 (2.4) | -0.02 | 4.1 (2.5) | -0.18 |
| Generous / Selfish | 3.3 (1.8) | 0.13 | 3.5 (2.1) | -0.07 | 3.4 (2.0) | 0.05 |
| Worthwhile / Worthless | 4.5 (2.0) | -0.05 | 4.3 (2.3) | 0.10 | 4.6 (2.5) | 0.07 |
| Interesting / Boring | 4.8 (2.4) | -0.15 | 4.5 (2.4) | -0.13 | 4.2 (2.4) | -0.27 |
| Knowledgeable / Ignorant | 3.9 (2.1) | 0.05 | 4.0 (2.0) | -0.18 | 3.6 (2.0) | -0.12 |
| Good / Bad | 4.2 (2.5) | -0.12 | 4.0 (2.5) | -0.02 | 3.9 (2.5) | -0.14 |
| BSES Total Score | 87.8 (28.0) | -0.09 | 85.2 (30.2) | -0.12 | 81.5 (33.8) | -0.21 |

Note: Participants indicated their agreement along a Likert scale for each pair of terms as applied to themselves. Higher scores represent more agreement with the *negative* term, i.e., *lower* self-esteem. For each timepoint, the mean score with deviation is shown. Effect sizes are calculated with the formula for Cohen's d. *BSES* – *Beck Self Esteem Scale*.

With regard toward attitudes toward goal-oriented tasks, participants reported increased intention to complete the tasks (Cohen’s d = 0.13) as well as increased confidence in their ability to do so (Cohen’s d = 0.35) after using a single session of NST-SPARK during Visit 1 (Table 5). At 1-week follow-up, the overwhelming majority of participants (90%) reported making progress in at least one of the tasks they had identified.

**Table 5:**
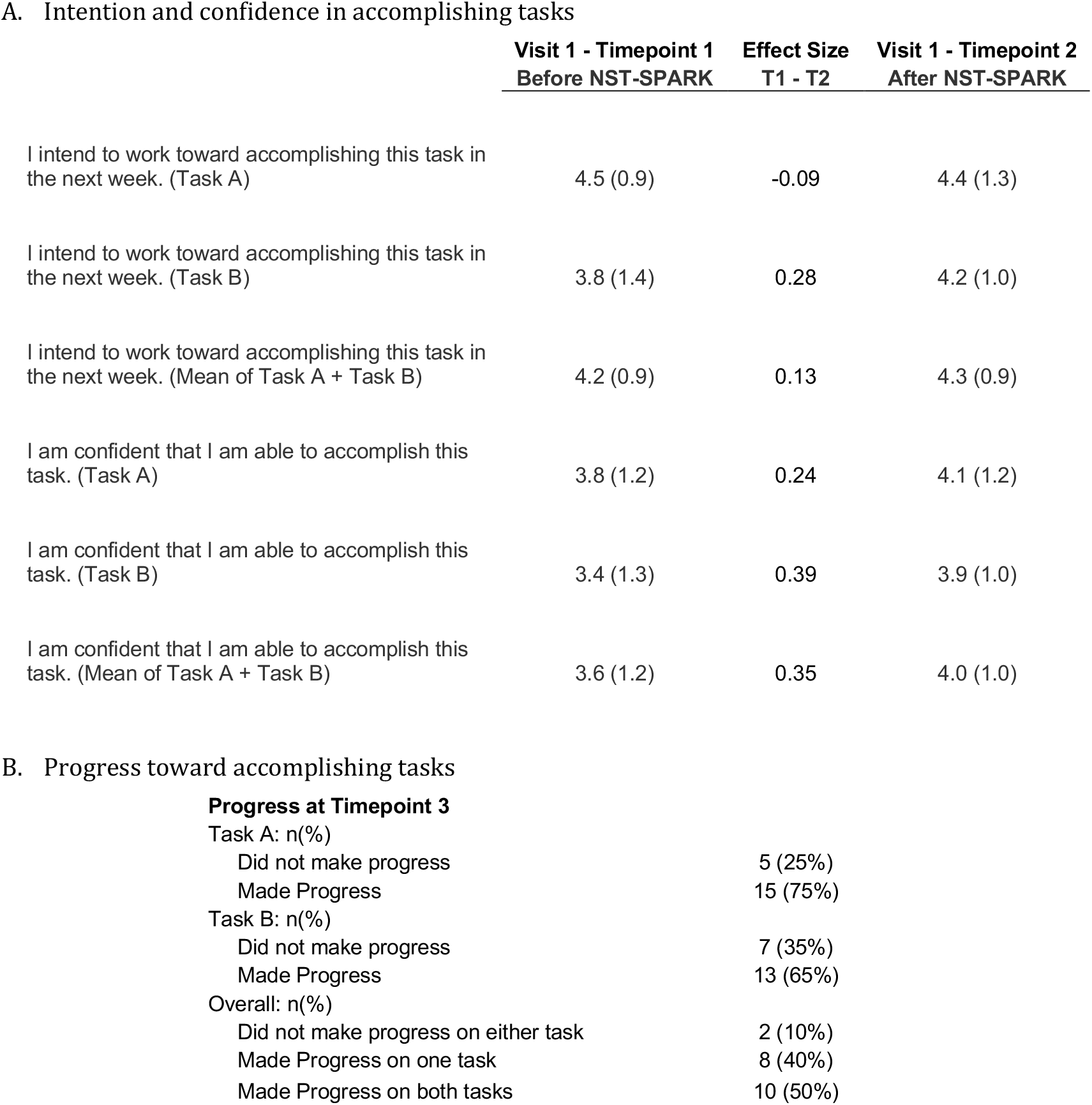
Attitudes toward goal-oriented activities.

**A. Intention and confidence in accomplishing tasks**
|  | **Visit 1 - Timepoint 1**<br>**Before NST-SPARK** | **Effect Size**<br>**T1 - T2** | **Visit 1 - Timepoint 2**<br>**After NST-SPARK** |
| --- | --- | --- | --- |
| I intend to work toward accomplishing this task in the next week. (Task A) | 4.5 (0.9) | -0.09 | 4.4 (1.3) |
| I intend to work toward accomplishing this task in the next week. (Task B) | 3.8 (1.4) | 0.28 | 4.2 (1.0) |
| I intend to work toward accomplishing this task in the next week. (Mean of Task A + Task B) | 4.2 (0.9) | 0.13 | 4.3 (0.9) |
| I am confident that I am able to accomplish this task. (Task A) | 3.8 (1.2) | 0.24 | 4.1 (1.2) |
| I am confident that I am able to accomplish this task. (Task B) | 3.4 (1.3) | 0.39 | 3.9 (1.0) |
| I am confident that I am able to accomplish this task. (Mean of Task A + Task B) | 3.6 (1.2) | 0.35 | 4.0 (1.0) |

## Discussion

### Key Findings

NST-SPARK v.1.5 is a single-module prototype for an AR app that employs gamification and the therapeutic principles of recovery-oriented cognitive therapy to target negative symptoms in schizophrenia. The primary objective of this study was to establish its acceptability and feasibility, and to inform further development of the app, if warranted. Secondary objectives were to explore changes in the targets of the intervention: defeatist beliefs, self-esteem (i.e., beliefs about the self), and attitudes and progress toward goal-oriented tasks.

In general, the goals of the prototype were achieved. Participants responded favorably to the app, overall, and particularly to the AR and gamification approach, as well as the therapeutic goal of reducing defeatist beliefs and avolition. They also used the app during the intervening 1-week period despite not being provided with additional incentives to do so. The main limitations identified had to do with the operation of the app itself and features not working as intended. We believe that these limitations can be readily addressed with further investment in a more complete version of NST-SPARK.

Additionally, while we cannot draw causal relationships from NST-SPARK due to the lack of a control group for this preliminary study, we did observe changes in defeatist beliefs, self-esteem, and goal-oriented tasks that were in the intended and hoped-for directions. This supports, though does not prove, the premise of NST-SPARK that it would target defeatist beliefs related to negative symptoms. Remarkably, 90% of participants made concrete progress toward at least one goal that they had identified during the intervention.

To our knowledge, there are no other apps using immersive technology to deliver a treatment for negative symptoms of schizophrenia. While there are other clinical trials underway, we are not aware of any published studies to which NST-SPARK can be compared.

### Future Considerations

The single-arm, unblinded design of this study is associated with obvious limitations but was a critical step in the design process to provide impetus for further development of NST-SPARK. Feedback obtained from the participants will be incorporated along with active engagement from individuals with lived experience with psychosis to produce a NST-SPARK v.2.0 that will deliver the therapeutic in a more complete form. This next iteration will be tested in a randomized controlled trial over a more extended period, and the effects on negative symptoms will be observed. The participants were relatively diverse with regard to race, ethnicity, gender, and socioeconomic background, including some participants on Medicaid/Medicare and those in community mental health programs. However, because this study was conducted remotely, the participants in this study may be enriched for individuals who had greater access to technology than average. Future studies will also include recruitment directly from treatment centers and efforts will be made to accommodate participants who may have lower access to technology.

## Conclusions

NST-SPARK is a mobile phone application that uses gamified AR experiences to delivery recovery-oriented cognitive therapy (CT-R). This preliminary, single-arm, unblinded study of a single-module prototype found that this approach is generally acceptable and feasible for people with SSD who have negative symptoms. Engagement of the intended target of defeatist beliefs was supported by our findings but require confirmation in future randomized controlled trials. Overall, we find that NST-SPARK is based on a promising approach and further development is warranted.

## Acknowledgements

We thank the participants for their time and willingness to provide feedback. This study was funded by North Shore Therapeutics, which is the creator of NST-SPARK.

## Conflicts of Interest

SXT, APB, MLB, SB, LMB, and JMK are affiliated as advisors and consultants for North Shore Therapeutics and are also employed full-time by academic institutions as described above. North Shore Therapeutics does not have any formal relationship with any of the above academic institutions. EY and WC are full-time employees of North Shore Therapeutics. In addition, SXT received research funding and serves as a consultant for Winterlight Labs, is on the advisory board and owns equity for Psyrin, and serves as a consultant for Catholic Charities Neighborhood Services and LB Pharmaceuticals. JMK has received consulting fees or Honoria for lectures from Alkermes, Boerhinger Ingelheim, Cerevel, Click Therapeutics, Intracellular Therapies, H. Lundbeck, HLS, Janssen, Johnson and Johnson, Merck, Minerva, Neurocrine, Newron, Otsuka, Roche, Saladax and Teva. He is a shareholder in Cerevel, HealthRyhthms, LB Pharma, North Shore Therapeutics and The Vanguard Research Group. He has received grant support from H. Lundbeck, Otsuka, Merck, Sunovion and Valera.

### Abbreviations

SSD: schizophrenia spectrum disorder
NST: North Shore Therapeutics
FDA: United States Food and Drug Administration
AR: augmented reality
VR: virtual reality
CT-R: recovery-oriented cognitive therapy
SCID-5: Structured Clinical Interview for The Diagnostic and Statistical Manual of Mental Disorders, Fifth Edition
SANS: Scale for the Assessment of Negative Symptoms
DBS: Defeatist Belief Scale
BSES: Beck Self-Esteem Scale
ATT: Attitudes Toward Behavioral Change
AIM: Acceptability of Intervention Measure
FIM: Feasibility of Intervention Measure

## Data Availability

Deidentified raw data and R code for data processing and analyses are available on our GitHub repository (https://github.com/NST-Spark/NST_Spark1b).

## Appendix A Attitudes Toward Behavioral Change (NST-ATT)

(Likert scale applied to agree/disagree questions below.)

DEFINITELY DISAGREE NEITHER AGREE/DISAGREE DEFINITELY AGREE

1 2 3 4 5

### Part 1 (before NST-SPARK)

1. Is there anything on your to-do list currently, any projects you would like to work on, or anything you would like to accomplish in the near future?
  a. (Probe as needed. If not much content) Is there anything that your family or friends have been encouraging you to do, but that you haven’t done?
  b. (If no response or unsure – ask about chores, meeting up with a friend or family member, improving health, getting a job or going to school.)
2. Which of these tasks seems the easiest to tackle? TASK A
3. Which of these is the most important to you? TASK B (get a different response if they initially answer the same task).
4. Thinking about TASK A, how much would you agree or disagree with the following?
  a. I intend to work toward accomplishing this task in the next week.
  b. I am confident that I am able to accomplish this task.
5. Thinking about TASK B?, how much would you agree or disagree with the following?
  a. I intend to work toward accomplishing this task in the next week.
  b. I am confident that I am able to accomplish this task.

### Part 2 (after NST-SPARK)

1. Before, you said that TASK A was something you would like to accomplish. How much would you agree or disagree with the following, now?
  a. I intend to work toward accomplishing this task in the next week.
  b. I am confident that I am able to accomplish this task.
2. What would be the first step to accomplishing TASK A? (Guide the participant to a concrete small step, e.g. put all the dirty clothes in the hamper; sign up for a handshake account; make a grocery list)
3. Before, you said that TASK B was something you would like to accomplish. How much would you agree or disagree with the following, now?
  a. I intend to work toward accomplishing this task in the next week.
  b. I am confident that I am able to accomplish this task.
4. What would be the first step to accomplishing TASK B?

### Part 3 (1wk follow-up)

1. Last time, you said that TASK A was something you would like to accomplish. Did you make any progress on it over the past week? YES/NO (description)
2. Last time, you said that TASK B was something you would like to accomplish. Did you make any progress on it over the past week? YES/NO (description)

## Appendix B: NST-SPARK Semi-Structured Feedback Interview

Assessor/Moderator to elaborate or ask follow up questions as needed.

### 1. Visit 1

a. What are your first impressions?
b. What do you like most about the app?
c. What do you like least about the app or what are some areas that need to be improved?
d. Did you experience any negative effects from using the app? For example, dizziness, vertigo, or changes in your symptoms?
e. Do you think you’d be able to use this app from a technical perspective? Does it seem simple enough to operate? If no, which parts seemed confusing or difficult?
f. (Imagine that we added similar but more elaborate experiences for a 12-week long experience, and also provided guides for reaching your goals in the real world.) Would you use this app if your doctor prescribed it for you? What makes you say that?
g. If there were other activities like the one you saw, what would make it fun and engaging for you?
h. Is there any other feedback that you’d like to provide to help us make this app better?

### 2. Visit 2

a. Did you get a chance to use the app during the week? If yes, how was your experience? If no, what got in the way?
b. Did you tell anyone about the app? Show it to friends or family?
c. Did you experience any negative effects from using the app? For example, dizziness, vertigo, or changes in your symptoms?
d. (Imagine that we added similar but more elaborate experiences for a 12-week long experience, and also provided guides for reaching your goals in the real world.) Would you use this app if your doctor prescribed it for you? What makes you say that? If yes, how often do you think you would use it on a weekly basis?
e. Do you have any more thoughts about what would make it fun and engaging for you?
f. Is there any other feedback that you’d like to provide to help us make this app better?

